# Participatory syndromic surveillance as a tool for tracking COVID-19 in Bangladesh

**DOI:** 10.1101/2020.08.28.20183905

**Authors:** Ayesha S. Mahmud, Shayan Chowdhury, Kawsar Hossain Sojib, Anir Chowdhury, Md. Tanvir Quader, Sangita Paul, Md. Sheikh Saidy, Ramiz Uddin, Kenth Engø-Monsen, Caroline Buckee

**Affiliations:** Department of Demography, University of California, Berkeley, USA; a2i Programme, Bangladesh; Telenor Research, Telenor Group, Norway; Center for Communicable Disease Dynamics, Department of Epidemiology, Harvard TH Chan School of Public Health, Boston, USA

**Keywords:** COVID-19, SARS-CoV-2, Surveillance, early-warning

## Abstract

Limitations in laboratory diagnostic capacity and reporting delays have hampered efforts to mitigate and control the ongoing coronavirus disease 2019 (COVID-19) pandemic globally. To augment traditional lab and hospital-based surveillance, Bangladesh established a participatory surveillance system for the public to self-report symptoms consistent with COVID-19 through multiple channels. Here, we report on the use of this system, which received over 3 million responses within two months, for tracking the COVID-19 outbreak in Bangladesh. Although we observe considerable noise in the data and initial volatility in the use of the different reporting mechanisms, the self-reported syndromic data exhibits a strong association with lab-confirmed cases at a local scale. Moreover, the syndromic data also suggests an earlier spread of the outbreak across Bangladesh than is evident from the confirmed case counts, consistent with predicted spread of the outbreak based on population mobility data. Our results highlight the usefulness of participatory syndromic surveillance for mapping disease burden generally, and particularly during the initial phases of an emerging outbreak.

## 1. Introduction

The ongoing coronavirus disease 2019 (COVID-19) pandemic has overwhelmed the healthcare systems of countries around the world, exposing the challenges faced by public health agencies when responding to rapidly emerging outbreaks. In particular, the scarcity of reliable data on the incidence of COVID-19 cases has hindered a timely response. On a national scale, control efforts should be guided by accurate data on cases and disease burden, ideally captured through widespread surveillance. However, very few countries affected by COVID-19 have sufficient viral testing capacity to monitor cases occurring in the community adequately. Hospitalization and death rates provide relatively robust indicators of SARS-CoV-2 transmission in some areas, but these are lagged by about 2 and 3 weeks, respectively. Identifying alternative indicators of transmission that reflect the timing of new infections is therefore an important priority for responding to the epidemic.

The first COVID-19 confirmed case occurred in Bangladesh on March 8th, with nearly 200,000 confirmed cases by July 15. SARS-CoV-2 testing capacity has increased significantly from a daily average of fewer than 100 tests in March to about 15,000 in June. However, as in most countries, the testing capacity can only cover a small fraction of even symptomatic cases. Reporting delays in rural and remote parts of the country also make it difficult to monitor the epidemic across the country in real-time. To augment surveillance, a participatory surveillance system based on self-reported symptoms via national telephone hotlines and the internet, assisted by a telemedicine team of clinicians, was deployed in March and rapidly scaled up over the course of the first few months of the outbreak.

The participatory surveillance system was set up through a public-private partnership, and is designed to collect syndromic information, to identify potential disease hotspots, and to provide information about COVID-19 to participants. Any surveillance data that relies on self-reported symptoms to monitor transmission will be subject to a range of biases, including the extent to which people are aware of and know how to use the system, and reporting behavior of people in the middle of a pandemic, which has naturally created much fear and uncertainty. Given the lack of specificity of the main symptoms of COVID-19, namely fever and cough, we also expect many people experiencing symptoms to have another disease unrelated to the coronavirus outbreak. Nevertheless, an uptick in individuals reporting symptoms consistent with COVID-19, particularly if verified through an interview with a clinician, may provide important insights into transmission hotspots.

While participatory crowdsourced syndromic surveillance has been utilized in many contexts [1]–[7], including for COVID-19 [8], [9], their ability to track an emerging outbreak at a high spatial resolution has not been evaluated previously. Here, we show that one such system, though noisy, provides an indication of where and when to expect new cases, suggesting that it could be a useful model in other places that need to map COVID-19 risk for decision making. The syndromic data suggests that the outbreak had spread across the country much faster than is evident from official case counts, consistent with geographic spread based on population mobility data. This system was developed rapidly, and we emphasize that the methods used to analyze the data are simple, reflecting both the urgency of establishing surveillance and the ease and reproducibility of the analysis. Nevertheless, we believe this is a useful, generalizable approach for participatory surveillance during an emerging epidemic.

## 2. Methods

### Data

The self-reported syndromic surveillance data analyzed here was compiled from three main sources: 1) a hotline number equipped with an Interactive Voice Response (IVR) system, 2) several internet and mobile applications and 3) an unstructured supplementary service data (USSD) based messaging system (see Figure 1). All systems are available free of charge to the user. Individuals are asked to report on their symptoms (cough, fever, and shortness of breath), contact with people with symptoms, contact with someone who had tested positive for COVID-19, and in some cases, their age and gender (see Supplementary Methods S1 for additional detail). Each response in the system is geolocated based on the nearest cell phone tower of the respondent, and mapped to *upazilas*, the operationally relevant administrative units in Bangladesh.

**Figure 1:**
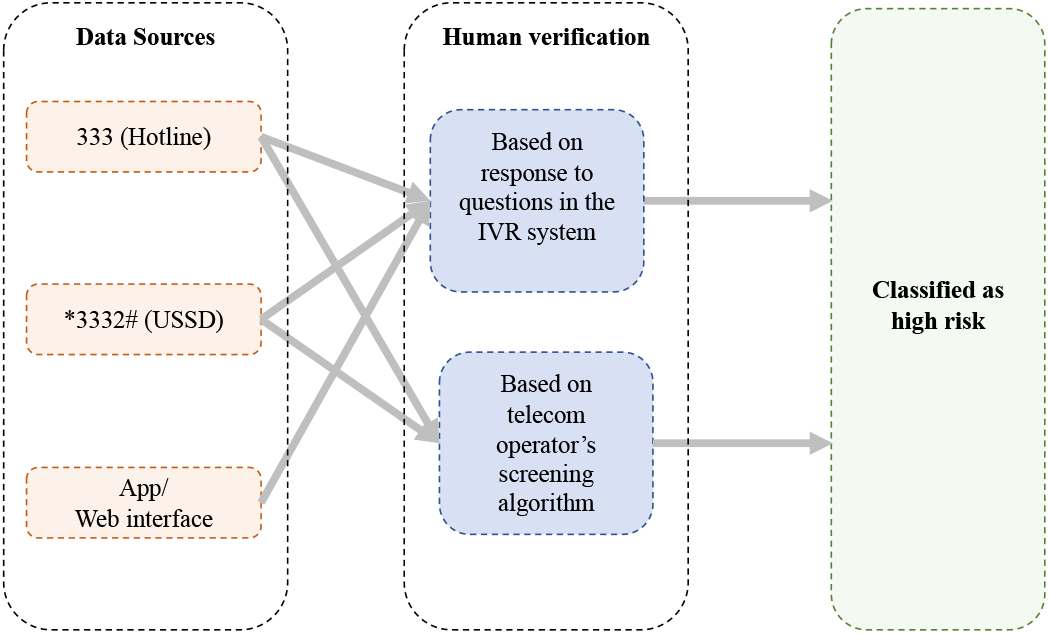
Self-reported syndromic data streams and human verification process

A subset of individuals reporting symptoms is connected with a human verifier – telemedicine doctors, healthcare professionals or trained field workers – for a preliminary diagnosis, based on their responses to the questions and algorithms specific to each mobile phone operator (see Supplementary Methods S1 for additional detail). Following an evaluation over the phone with a human verifier, individuals are then classified as having high or low risk for COVID-19. This classification is based on the reported symptoms, contact with people with symptoms, and, to some extent, the judgement of the human verifier. In the initial phase of the outbreak, high risk classification was also based on whether or not the individual was from an area with confirmed COVID-19 cases. Hence, we see a stronger correlation of the number classified as high risk with reported cases at the start of the outbreak (Supplementary Figure S1). Since self-reported data is inherently noisy, here we focus on the subset of people reporting multiple symptoms consistent with COVID-19 and those who were connected with a human verifier.

We combine these data from the multiple sources to establish i) the number of individuals reporting multiple symptoms consistent with COVID-19 and ii) the number of suspected cases that are classified as high risk following human verification. We estimate these values for each of the 544 upazilas. To adjust for variations in population density in different upazilas, we calculate both the raw counts as well as the per capita count for each metric using population data from *Worldpop* [10]. The self-reported syndromic surveillance system was ramped up and widely advertised starting at the beginning of April. Here, we analyze data available from April 1 to June 15. To reduce noise in the daily data, we sum the measures by week. Results are qualitatively similar when using two-week windows.

### Validation of syndromic data with confirmed case counts

While testing capacity is limited and spatially heterogeneous, in the absence of other robust indicators it is currently used as the key epidemiological indicator guiding decision-making in Bangladesh. To establish how well the self-reported data reflects at least this imperfect signal of the epidemic, we compare the trends in the number reporting symptoms and those classified as high risk cases – which we assume reflect symptom onset – to lab-confirmed cases with varying time lags, to account for the delay in lab-confirmed testing and reporting. In the early phase of the outbreak, before testing capacity was expanded, individuals were identified for testing through the human verification process. Again, this can explain the stronger correlation of the number classified as high risk with confirmed cases at the start of the outbreak (Supplementary Figure S1). The syndromic surveillance system is not currently used to inform testing locations, so would not influence the association between confirmed cases and self-reported data.

We calculated correlations between lab-confirmed cases and each of the syndromic indicators in an upazila for every week separately to understand potential changes in the association over time (Supplementary Figures S1 and S2). We also fitted a linear mixed-effects model to estimate the relationship between the syndromic indicators and confirmed cases over time, with a random effect to capture upazila-level differences. Specifically, we modeled the number of confirmed cases in each upazila in each week as a linear combination of the syndromic indicator, the lagged value of the syndromic measure at one and two weeks, and a random effect for each upazila. We estimated separate models for the two syndromic indicators (see Supplementary Methods S2 for full model specification).

### Comparison with predicted importation based on population mobility

Since test capacity was especially limited at the start of the outbreak, particularly in places outside Dhaka, we also compare the syndromic data to predictions of imported cases from Dhaka to the rest of the country. The predictions are similar to those based on a Chikungunya outbreak in 2017 [11] and are based on population mobility estimates from mobile phone data and estimated prevalence at the start of the outbreak assuming a simple exponential growth model (Supplementary Methods S3). We use previously described methods to construct population mobility estimates from mobile phone call detail records (CDR) from the largest mobile phone operator in Bangladesh, with over 64 million subscribers. Here, we use CDR data available for April 2017 as an estimate for the pre-lockdown movement patterns in Bangladesh. To test the validity of this assumption, we compare the CDR data for April 2017 to CDR data available for May 2020. We find a strong correlation between the proportion of travelers traveling from Dhaka to all other upazilas in 2017 and in 2020 (Supplementary Figure S3). Since the CDR data for 2020 is unavailable for the pre-lockdown business-as-usual period in March, we use the 2017 mobility estimates to derive the relative distribution of travelers from Dhaka to each upazila outside Dhaka district, and adjust the volume of travel with the 2020 population estimate for Dhaka. We estimate the probability of importing at least one infected person, using the mobility estimates and an estimated prevalence for Dhaka at the start of the outbreak. Using previously described methods for the spread of COVID-19 [12], we estimate the probability of importing one or more infected cases in the first 16 days of the outbreak (i.e. prior to any travel restrictions) for each upazila outside Dhaka district.

## 3. Results

The self-reported syndromic surveillance system received nearly 3.5 million responses between April 1 and June 15. There is considerable temporal variation in the total response volume across the three systems. For instance, a large spike occurred in the responses to the USSD system in April due to targeted messaging from the four mobile phone operators (Supplementary Figure S4). Despite the noise and initial volatility, the number of people reporting symptoms and the number classified as high risk following human verification exhibits a general increasing trend in line with the epidemic trajectory in Bangladesh (Figure 2).

**Figure 2:**
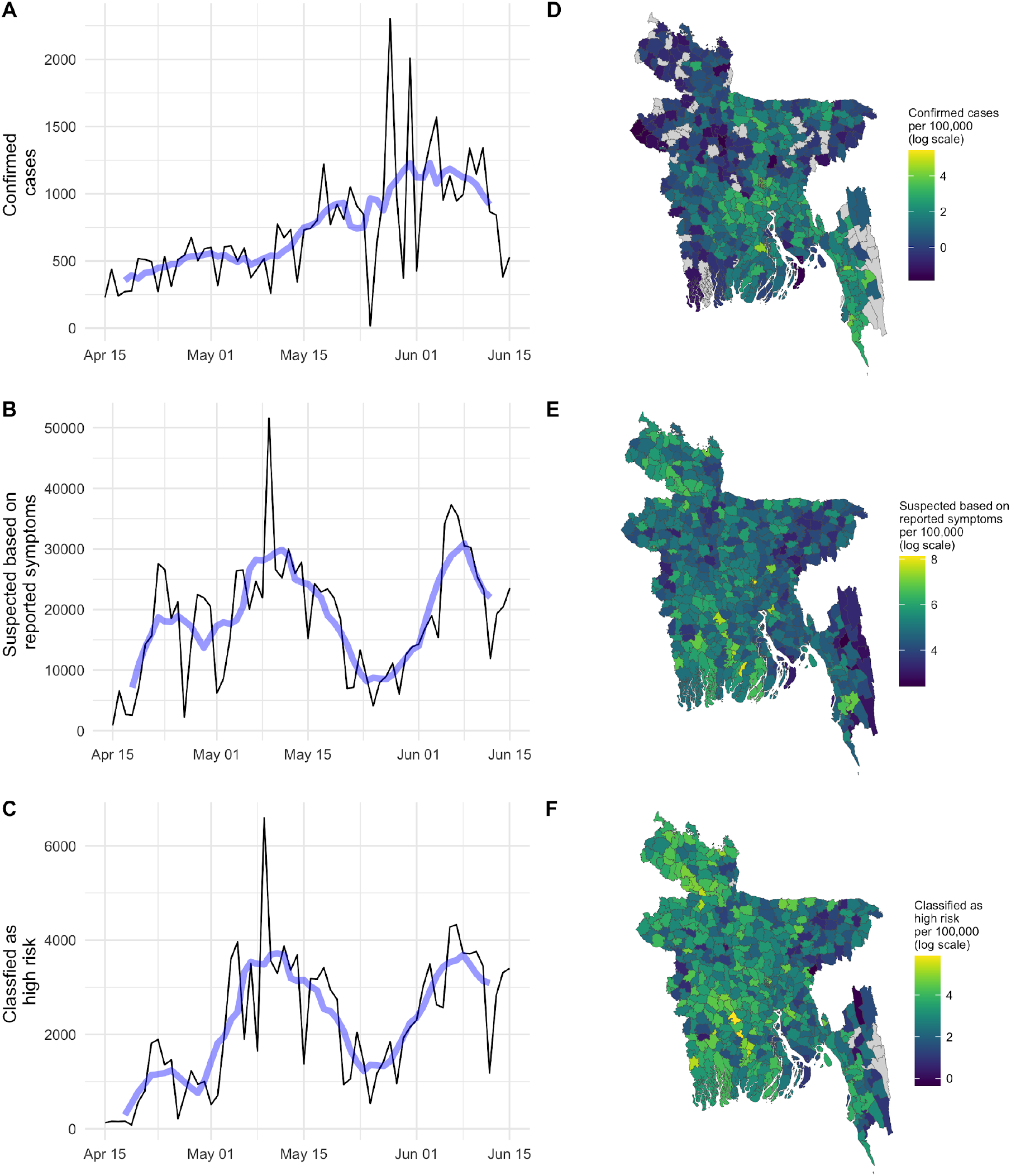
Timeseries of (A) daily number of confirmed cases, (B) total number classified as a suspected COVID-19 case and (C) the total number classified as being high risk of COVID-19 following human verification (bottom) from April 15 to June 15. Data prior to April 15 are not comparable as several of the systems were not set up until April 15 and are, thus, not shown here. The blue line shows the seven-day rolling average. The spatial distributions are shown in (D), (E), and (F) respectively. The heatmaps show each measure summed over a two-week window from June 1 to June 15 (on a log scale).

### Association with confirmed case counts

The syndromic surveillance data shows strong associations with confirmed cases over the same time period and with a lag time of one to two weeks (Figures 2 and 3). A spike in the number of people reporting symptoms, and those considered high risk, occurred nationally during the first few weeks of May, and a spike in lab confirmed cases followed at the end of the month. The erratic pattern of confirmed cases at the end of May is most likely a result of the major Eid holidays, during which both testing and the reporting of test results may have been delayed. Interestingly, there is also a decline in the self-reported numbers during the Eid holidays, perhaps reflecting a lower propensity of reporting symptoms during the holiday period.

**Figure 3:**
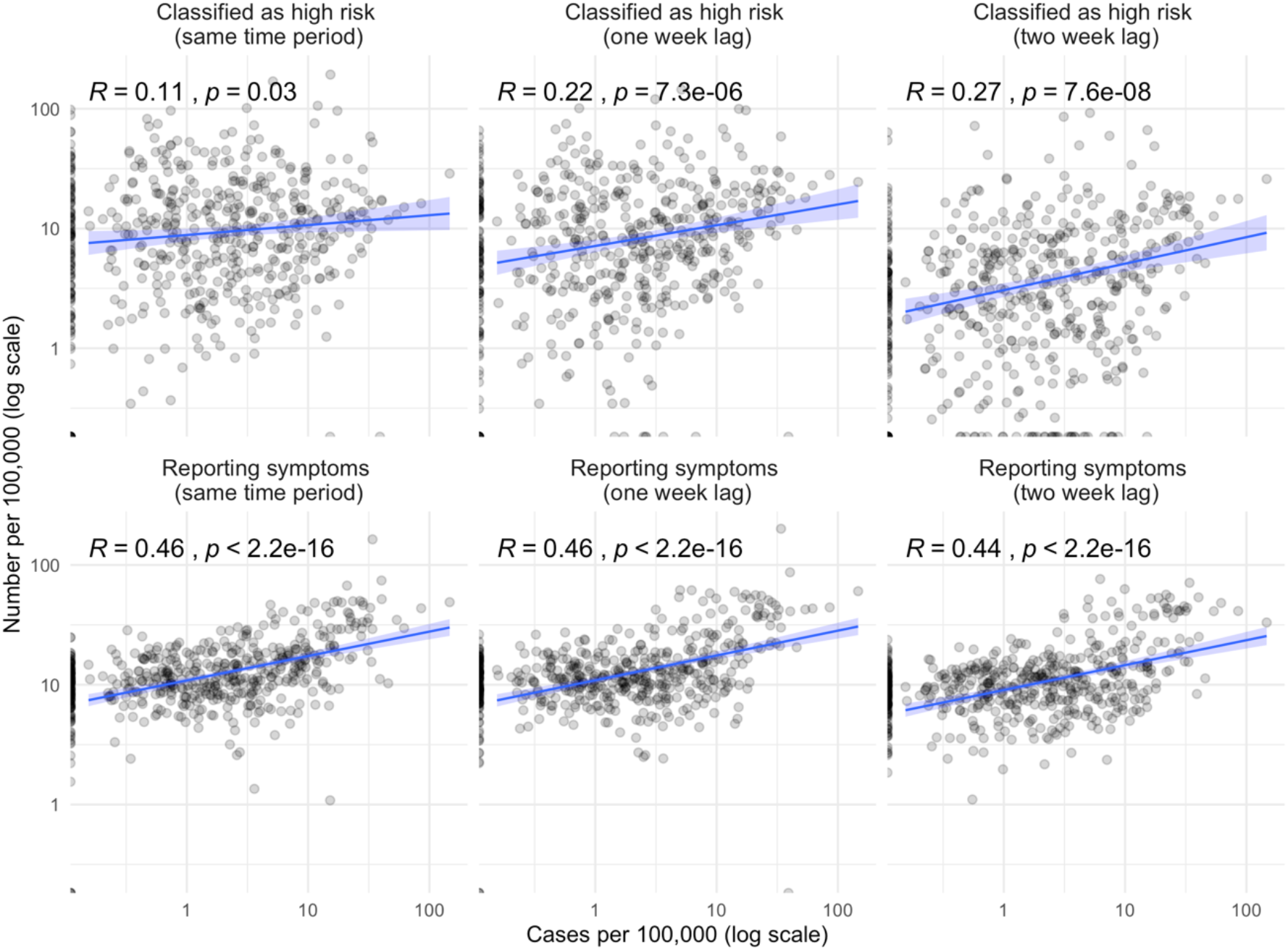
Top row: Correlation between confirmed cases per 100,000 and the number of people who are classified as high risk for COVID-19 following human verification (summed over a one week window from June 8 to June 14) for the same time period, with a one week lag, and a two week lag. Bottom row: Correlation between confirmed cases per 100,000 and the number of people reporting symptoms consistent with COVID-19 infection (summed over a one-week window from June 8 to June 14) for the same time period, with a one-week lag, and a two-week lag. All plots show the Pearson correlation coefficient for the bivariate relationship and associated p-values.

The number of people reporting symptoms consistent with COVID-19 and the number classified as high risk are both significantly correlated with observed cases in the same time period, as well as at lags of one and two weeks. The correlation for the most recent seven days (June 8 to June 15) is shown in Figure 3 (correlations for each week is shown in Supplementary Figures S1 and S2). When we compare the timeseries and include a random effect to account for upazila-level differences, such as population size, we find the largest positive association between confirmed cases and the syndromic data in the same time period followed by lags of one week and two weeks (Figure S5). The observed positive relationship suggests that the selfreport system may be a useful indicator of an uptick in cases in particular regions as the epidemic unfolds. We see a stronger association between high risk cases and confirmed cases at the start of the outbreak (likely due to reasons described above), and, interestingly, a stronger association between the number reporting symptoms and confirmed cases in more recent time periods (Figure S2). Monitoring and evaluating these temporal changes will be crucial for assessing the reliability of self-reported data as the pandemic evolves.

### Early spread of COVID-19 in Bangladesh

Data from the syndromic surveillance suggests that the outbreak was much more widespread at the start of April, about a month after the first confirmed case, than is evident from the data on confirmed cases. Confirmed case counts remained low in the month following the first detected case on March 8, reflecting the very limited testing capacity at the start of the outbreak. Initial testing was limited to travelers arriving from abroad and known contacts of confirmed cases. Until March 27 a single laboratory was responsible for administering and analyzing COVID-19 tests in Bangladesh, and only about 1600 total samples were tested nationally by the end of March [13]. Testing was also largely limited to Dhaka until the end of March [14]—[16].

While the number of laboratory-confirmed cases remained low in March and April, even as early as April 1 (when the syndromic surveillance system was not yet fully set up), nearly all upazilas had individuals reporting symptoms consistent with COVID-19. The vast majority of upazilas had individuals reporting symptoms as well as individuals classified as being at high risk for COVID-19 before the first confirmed case was reported (Figure 5), with a median lead time of 10 days. Confirmed cases in many upazilas were first reported at the beginning of April, and cases across the country began to increase from mid-April onwards, reflecting the increase in testing capacity and the expansion of testing beyond Dhaka.

**Figure 5:**
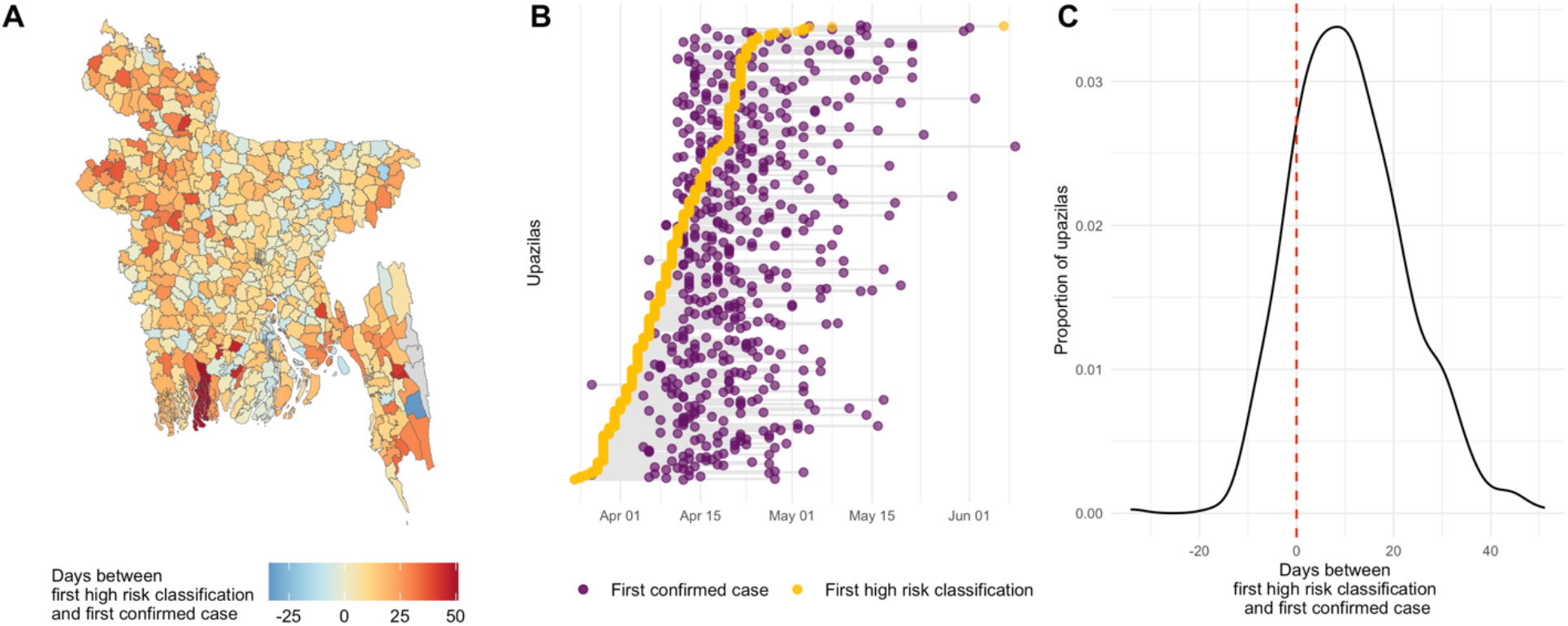
Days between the first confirmed case and the first high risk classification based on the self-reported syndromic surveillance. Spatial variation in the time between the first confirmed case and the first high risk classification shown in (A) and the respective dates by upazila are shown in (B). Upazilas are ordered according to the date of their first high risk classification in (B). (C) shows the density plot for the lag between first high risk classification and first confirmed case. Red line indicates zero days of lag time. A positive lag time indicates that the first high risk classification was reported before the first confirmed case. The median lag time across all upazilas was 10 days.

**Figure 6:**
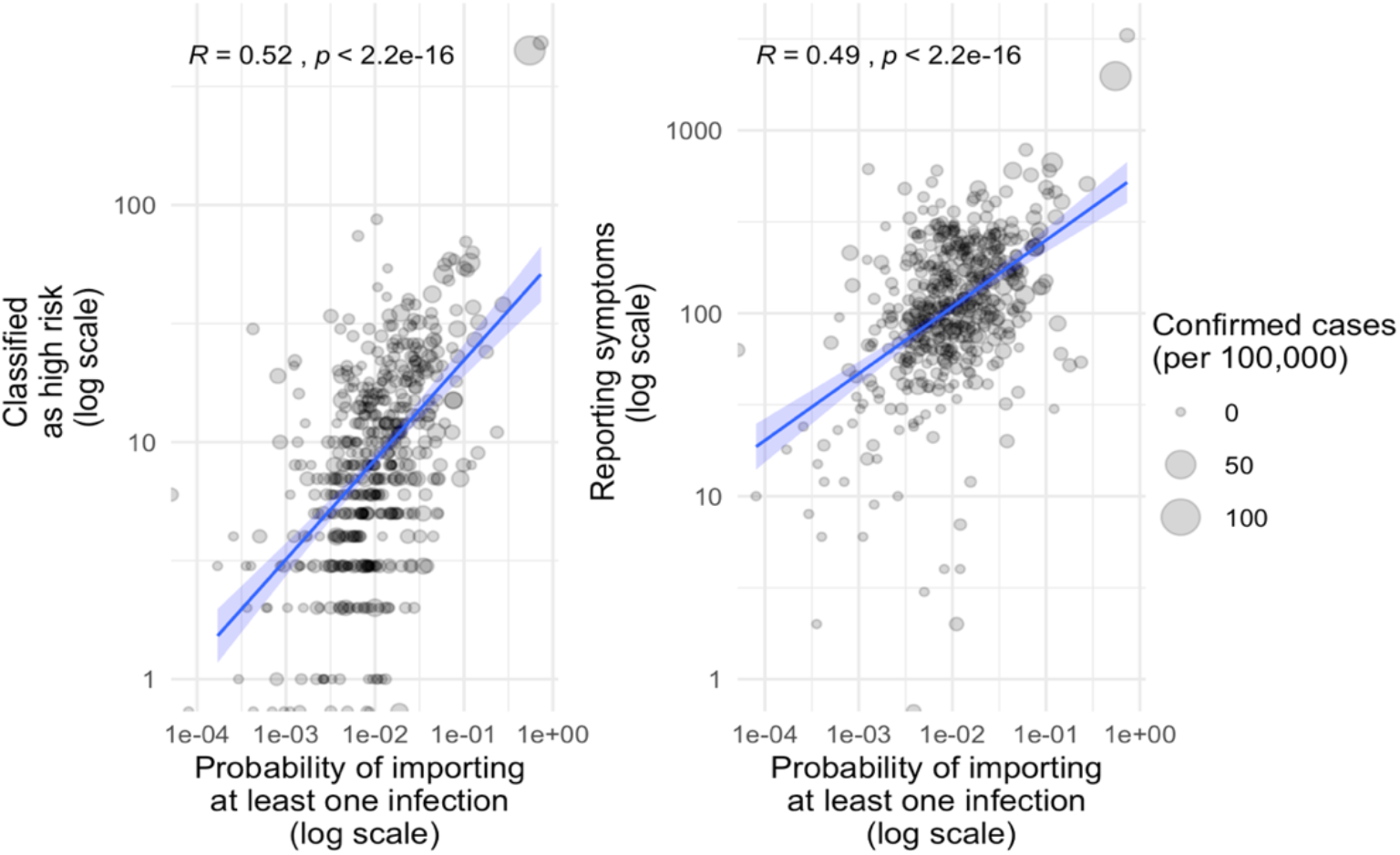
Correlation between syndromic data (for first two weeks that data from all surveillance streams is available: April 15 to April 28) and probability of importation estimated using mobile phone data. Both plots show the Pearson correlation coefficient for the bivariate relationship and associated p-values.

The early spread of the outbreak suggested by the syndromic surveillance is also consistent with the predicted importation across the country based on population mobility data. We find a significant positive association between the probability of an upazila having an imported case by March 19, before the lockdown and physical distancing measures were put in place, and the syndromic surveillance indicators. Taken together, these results are indicative of an early spread of the outbreak, which is not captured in the official case counts. These results also demonstrate the potential usefulness of participatory surveillance as an early-warning system during the initial phases of an emerging outbreak.

## 4. Discussion

The ongoing COVID-19 pandemic has highlighted the limitations of traditional surveillance systems in terms of their timeliness and scalability. Unlike traditional surveillance that requires patients to interact with the healthcare system, and are limited by testing capacity and reporting delays, participatory surveillance relies on the self-reporting of symptoms. Crowdsourced participatory surveillance, through phone hotlines, mobile phone applications, and the internet, have been used in many contexts and have shown the greatest promise for influenza surveillance [7], [17]-[20]. Given the rapid global increase in the use of mobile phones and access to the internet, these surveillance systems can provide a useful complement for more traditional surveillance systems [2].

Here, we show the potential use of participatory syndromic surveillance for tracking the COVID-19 outbreak in Bangladesh. These self-reported systems were set up very rapidly as the COVID-19 epidemic started to emerge in Bangladesh. Although we observe considerable noise in the data and initial volatility in the use of the different reporting mechanisms, as expected, the self-reported data is positively correlated with confirmed cases at the upazila level a week later. Since the data from this tool is not being used to guide testing currently, we can be reasonably confident that the signal is not the cause of more testing, but it remains difficult to determine how tests are allocated across the country. We must therefore rely on confirmed cases as a way to establish the congruence of these indicators, but we acknowledge that there could be unobserved epidemic dynamics that remain hidden from both kinds of surveillance data.

The self-reported surveillance system also suggests an earlier spread of the outbreak across Bangladesh than is evident from the confirmed case counts. On average, the first high risk classification based on the self-reported data preceded the first confirmed case by a little over a week. Nevertheless, we observed a more wave-like dynamic among the high-risk classified calls compared to confirmed cases, which emerged very rapidly across the country. We believe this is likely to reflect the roll-out of testing capacity, and that the self-report data offers reasonable insights into the early stage of the epidemic. The lead-time offered by this kind of system could be particularly important for monitoring reintroductions and potential resurgence across different parts of the country in the future. This system can also be useful for prioritizing the efforts of community support teams that are currently being deployed in parts of the country to increase awareness and promote non-pharmaceutical interventions.

The self-reported surveillance system is inherently noisy and subject to a range of reporting biases. Nevertheless, we believe this system provides a useful approach to augment surveillance for a disease like COVID-19, where testing capacity is limited, and symptoms are relatively non-specific. Ideally, this would be combined with other syndromic surveillance data from clinics and hospitals, as well as hospitalization and death rates that are COVID-19 specific. As the epidemic unfolds in Bangladesh, this system can be continuously validated and improved, and maintained in its aftermath as a more general public health surveillance system. By directly engaging with the population at risk, participatory surveillance also provides a channel for disseminating public health guidelines and information and can enable a more rapid response to public health emergencies.

## Data Availability

Data was made available through partnership with a2i in Bangladesh.

## Acknowledgements

The authors thank Directorate General of Health Services (DGHS), Bangladesh, and the Institute of Epidemiology Disease Control and Research (IEDCR) for providing data and for feedback on the analysis. The authors thank Grameenphone, Banglalink, Robi, Teletalk, the Bangladesh Telecommunication Regulatory Commission (BTRC), and the National Telecommunications Monitoring Centre (NTMC) for providing access to the telecommunications data; BRAC and CMED for community data; and Cramstack and Nascenia for data analysis. Research effort by COB was enabled by a Maximizing Investigator’s Research Award from the National Institute of General Medical Sciences (R35GM124715).

